# Design and evaluation of a youth co-designed trauma-informed public health resource for use in public sector settings in England

**DOI:** 10.64898/2026.08.05.26359401

**Authors:** Siobhan Hugh-Jones, Luke Allder, Ellie Baker, Isabelle Butcher, Harsimran Sansoy, Nicola Shaughnessy, Kamaldeep Bhui

**Affiliations:** School of Psychology, University of Leeds, UK; School of Arts and Architecture, University of Kent; Institute for Arts in Therapy and Education, London, UK; Research and innovation Services, University of East Anglia; Department of Psychiatry, University of Oxford

## Abstract

**Background:** Trauma-informed approaches (TIAs) are increasingly implemented across public-sector settings to improve support for young people affected by adverse childhood experiences (ACEs). However, practitioners often report difficulties translating broad trauma-informed principles into everyday practice, and young people are rarely involved in developing resources intended to support implementation.

**Aim:** To co-design, implement and undertake a preliminary evaluation of a youth-led trauma-informed resource for professionals working with young people in public-sector settings in England.

**Methods:** The study formed part of the UKRI-funded Attune programme and employed Accelerated Experience-Based Co-Design (AEBCD). Eighteen adolescents with lived experience of ACEs and 16 professionals from nine public-sector settings participated in three regional co-design workshops. Findings from a prior arts-based lived experience study informed the workshops. Participants collaboratively developed *Validating Voices*, a low-cost resource designed to increase validating interactions between professionals and young people. The resource was subsequently introduced into nine organisations and evaluated using staff surveys and semi-structured interviews.

**Results:** Co-design participants identified professional invalidation of young people’s experiences, identities, needs and emotions as an under-recognised contributor to mental health. The resulting resource combined discussion cards, creative activities, role-play and organisational reflection exercises to promote validating practices. Five organisations implemented the resource and reported it to be feasible. Flexible local adaptation was common, while more participatory role-play elements proved harder to implement consistently. Staff observed increased opportunities for disclosure, reflection, peer connection and professional curiosity about young people’s experiences. Staff reported listening differently to young people and, in some settings, implementing changes in response to young people’s recommendations.

**Conclusions:** Youth-led co-design identified validation as a practical and meaningful mechanism for operationalising trauma-informed principles in everyday professional practice. With refinements, *Validating Voices* shows promise as a resource to support more relational, collaborative and trauma-informed responses to young people in public sector settings.

## 1.0 Introduction

Adverse childhood experiences (ACEs) are highly stressful events that occur before age 18 encompassing various forms of abuse, neglect, household adversity and dysfunction, as well as community violence (Felliti et al.,1998). Decades of evidence demonstrate that ACEs are a significant potential determinant of mental health outcomes during adolescence (Obse et al. 2026; Sahle et al., 2022). This is partly because many ACEs include exposure to trauma, and the behavioural and emotional patterns that emerge in their aftermath are often understood as trauma responses (Friedman et al., 2011; van der Kolk, 2003). Such responses are diverse and may include heightened vigilance, difficulties in emotional regulation, a strong need for predictability, low self-worth and reduced trust in other (Bellis et al., 2023).

Although these patterns represent survival and adaptive responses to adversity, when manifested in adolescents and young people, they are frequently misinterpreted by adults as oppositional, unmotivated, or disruptive behaviour. In the UK and internationally, many settings where professionals routinely encounter adolescents (e.g. schools, social, health and mental health services, police) are unaware of how a young person’s trauma may be shaping their needs, engagement and behaviour (Rosenthal et al., 2016; Spence et al., 2021). Poor understanding and inadequate responses from adults pose further risks to adolescents’ mental health and their longer-term recovery from adversity and trauma.

Efforts to respond better to adolescents with ACEs increasingly centre on trauma-informed approaches (TIAs) (Champine et al., 2022; Sweeney et al., 2018; Yatchmenoff et al., 2017). Before proceeding, we acknowledge that terminology in this field remains contested and is used inconsistently across policy, research and practice. Terms such as trauma-informed, trauma-responsive, trauma-focused, and references to trauma-informed principles are sometimes used to describe overlapping, but not identical, approaches (Hanson et al., 2018). For the purposes of this paper, we use trauma-informed approach (TIA) as an umbrella term to refer to organisational and relational practices that recognise the prevalence and impacts of adversity and trauma and seek to promote safety, trust, collaboration, empowerment and choice (Harris & Fallot, 2001). This definition reflects current UK guidance (Office for Health Improvement and Disparities, 2022), and aligns with the focus of our study on supporting everyday professional practice rather than trauma-specific clinical interventions.

TIAs aim to enhance organisational and practitioner capacity to recognise that young people’s engagement, behaviour and coping may be shaped by past adversity or trauma, and to ensure that services minimise the risk of re-exposure to harmful experiences (Berring et al., 2024). Emphasising safety, trustworthiness, empowerment, collaboration, and choice, TIAs promote a shift from deficit- or problem-oriented interpretations of behaviour toward understanding of the potential legacy of trauma on a young person (Harris & Fallot, 2001). In principle, TIAs in human services should involve holistic and sustained improvements in organisational policy, procedures and relational dynamics with the people they aim to support (Fallot & Harris, 2008; Oral et al., 2020).

The expansion of TIAs globally is significant, and although conceptualisations of TIA and methodologies to evaluate them remains contested, evidence of the positive impact of TIAs on traumatised young people is growing (Berring et al., 2024; Bunting et al., 2019; Dorado et al., 2016; Fernandez et al., 2023; Wall, 2021; Wright, 2026). In the UK, the government has defined principles for TIAs (Office for Health Improvement and Disparities, 2022), and the implementation and evaluation of TIAs is emerging in multiple sectors in the country, including in healthcare (Emsley et al.2022), education (Boylan et al., 2023), criminal justice (Quigg et al., 2025), social care (Asmussen et al., 2022) and mental health (Currie & Lynch, 2026).

Yet despite the potential of TIAs to be helpful to trauma populations, implementing them across diverse human services remains challenging, across both clinical and community settings, including those for young people (Berring et al., 2024; Steinkopf et al., 2022). Barriers include a lack of consensus on what a TIA looks like for a youth-centered setting / service, the complexity of changing organisation cultures and practices, and the lack of tools to help frontline professionals deliver a TIA as routine in their encounters with young people (Avery et al., 2021). Although guidance and resources to implement a TIA have emerged (e.g., SAMSHA’s guide for TI approaches; traumainformedcare.chcs.org; Scottish Government National Trauma Transformation Programme; Thirkle et al., 2022) they are often viewed as resource-intensive (Spencer et al., 2026) and rarely involve young people’s voice in their development (Koslouski & Chafouleas, 2022). This is a significant omission, reminding us that key voices are still being excluded from defining what ACEs and trauma are, and how settings and services should be designed in response (Becker-

Blease, 2017; Prior et al., 2022). Furthermore, organisations and services, and participatory agendas, continue to call for practical, youth-focused exemplars and actionable recommendations for applying TIA principles (Flodgren et al., 2025; Yatchmenoff et al., 2017). To date, no trauma-informed public resource designed collaboratively with young people and public-health stakeholders has been co-designed, implemented, and evaluated in England. Co-design is one way to redressing the exclusion of youth voice in research and service development and can enhance the acceptability and effectiveness of interventions by embedding lived and living experience, problem understanding and preferred responses (McCabe et al., 2023; Oliveras et al, 2018; Veldmeijer et al., 2023).

### Study aims

The present study aimed to co-design, implement, and conduct a preliminary evaluation of a youth-led resource to support professionals in UK public-sector settings to adopt a TIA (see Hugh-Jones et al., 2024 for the study protocol). The work was nested within Attune (2021–2026), a national UK Research & Innovation funded programme examining risk and resilience pathways through which ACEs shape adolescent mental health (Bhui et al., 2026). Across multiple sites and working with diverse young people aged 10–24, Attune employed participatory, arts-based methods to elicit young people’s lived experiences of adversity and to identify opportunities to mitigate its effects on their mental health. Methods and findings from the lived experience workstream, on which this study draws, are reported in detail elsewhere (Pavarini et al., 2021; Hugh-Jones et al., 2026).

Attune was designed so that learning from the lived experience workstream would inform a subsequent co-design phase involving a new group of ACE exposed young people working together with public sector professionals from settings in which they were affiliated. This collaborative group was tasked with developing a low-cost, low-burden resource for professionals that responded to the lived experience of adolescents with ACEs reported in the earlier workstream. Adopting participatory, youth-centred methods and decision-making powers in the development of trauma-informed resources remains relatively rare for adolescent community contexts. This paper reports the methods and findings of our co-design process and early learning from its implementation in public sector settings. We report how young people conceptualised the challenges arising from early adversity, what risks shaped their subsequent mental health outcomes and where they located opportunities for professionals to improve their support to them. The research questions underpinning this study were:

1. Co-Design Stage: What do young people and stakeholders want in a co-designed resource to improve public sector capacity to work in trauma-informed ways?
2. Evaluation Stage: Is the co-designed resource acceptable, feasible, efficacious and how should it be refined for future implementation in the UK public sector?

## 2.0 Materials and Methods

### 2.1 Ethical Approvals

The co-design and evaluation stages were approved by the University of Oxford’s Research Ethics Committee (R71941/RE001) and the UK’s Health Research Authority (23/WM/0105).

### 2.2 Accelerated Experience-Based Co-Design (AEBCD)

#### 2.2.1 Recruitment and Participants

Purposive recruitment was used to recruit professionals and adolescents from diverse settings. As per our protocol, we aimed to recruit a minimum of six public sector settings whose remit included supporting vulnerable young people (e.g., educational institutions, community youth programs, youth offending teams, domestic violence teams). Recruitment was via the Attune project’s network and by direct approach to settings in study regions. We sought settings from Attune’s three English study sites (Cornwall, Kent and Yorkshire), representing rural, coastal and urban regions. To be eligible, settings had to be (i) motivated towards working in trauma-informed ways and (ii) willing to nominate at least one member of staff to attend three full day, regional co-design workshops in partnership with at least one adolescent from their setting. Settings were encouraged to invite participation by adolescents from underrepresented groups, including those with marginalised ethnicities and identities, neurodivergences and / or disabilities. We report elsewhere on the importance of reaching underserved adolescents in ACE research (Bhui et al., 2026). Adolescents were eligible to participate if they (i) had a history of ACEs; (ii) were able to attend three co-design workshops with parental consent for under 16s; (iii) could converse in English; and (iv) were not in crisis or in need of urgent care as judged by themselves, parent and accompanying professional. Following professional, individual and parental consents, adolescents were asked to complete an intake survey prior to the co-design workshops. The survey included a range of standardised measures on ACEs, trauma and mental health as well as questions about their identities and support needs for project participation. Survey completion was an invitation made to all adolescents taking part in the broader Attune project to understand the participating cohort. Full details of survey items are reported in Bhui et al., (2026).

We recruited 16 professionals and 18 adolescents from 9 settings across our three study regions. Professionals and young people came as dyads or small groups from each setting to work in partnership. These were: a domestic abuse organisation (n=2 professionals and 2=adolescents), an LGBTQ+ community groups (n=1 professional, n=1 adolescent), two community hubs in areas of deprivation (n=3 professionals, n=5 adolescents), a care leaver’s service (n=2 professionals, n=1adolescent), two university student counselling services (n=4 professionals, n=3 adolescents), a city college (n=2 professionals, n=3 adolescents), and a specialist education secondary school for (n=2 professionals and n=3 adolescents) for students with an Educational, Health and Care Plan and who have Social, Emotional and Mental Health (SEMH) as their primary need. Attending professionals were typically key workers or senior staff. Limited demographics of the 15 adolescent co-design participants contributors who opted to provide these details in the Attune survey are reported in Table 1.

**Table 1:** Demographics of adolescent contributors (n=15) to the co-design stage.

| **In Age Range (y)** | **Ethnicity** | **Neurodivergence or Neurodivergent Identities** |
| --- | --- | --- |
| 17-21 | Hispanic | None. |
| 17-21 | English, Welsh, Scottish, Northern Irish or British | Described themselves as Autistic* and to experience selective mutism. |
| 13-16 | English, Welsh, Scottish, Northern Irish or British | Described themselves as Autistic and to experience ADHD dyscalculia, learning disabilities. Diagnosed dyslexia. |
| 17-21 | Caribbean | None. |
| 13-16 | Pakistani | None. |
| 13-16 | Turkish | Described themselves as language impaired. Received a clinical diagnosis of autism and dyspraxia. |
| 13-16 | English, Welsh, Scottish, Northern Irish or British | None. |
| 17-21 | Pakistani | None. |
| 13-16 | English, Welsh, Scottish, Northern Irish or British | Described themselves as receiving a clinical diagnosis of ADHD and autism. |
| 17-21 | English, Welsh, Scottish, Northern Irish or British | Described themselves as experiencing dyspraxia and language impairment. Received a clinical diagnosis of ADHD and autism. |
| 17-21 | Caribbean | Received a clinical diagnosis of autism. |
| 17-21 | Gypsy or Irish Traveller | None. |
| 13-16 | Gypsy or Irish Traveller | None. |
| 17-21 | English, Welsh, Scottish, Northern Irish or British | Described themselves as Autistic. |
| 17-21 | Chinese | Described themselves as experiencing ADHD and a learning disability. Diagnosed dyscalculia and dyslexia. |

\*We report 'Autistic' where an adolescent offered an identity-first self-description. For clinical diagnoses, we refer to 'autism'.

Adolescent ages ranged from13-21y (mean age 17y) and they reported diverse ethnicities, gender identities (n=9 female, n=3 male, n=3 non-binary or gender fluid, n=1 prefer not to say), sexualities (n=8 heterosexual, 4=bi or pansexual, 1=lesbian, n=2 prefer not to say), neurodivergences and neurodivergent identities, and mental health diagnoses. All professionals stated their ethnicity as White British, and their self-reported knowledge of working in trauma-informed ways varied from highly experienced (n=8 experienced) to no knowledge (n=7).

#### 2.2.2 Accelerated Experience-Based Co-Design (AEBCD) Method

AEBCD is suitable for bringing collected narratives of experience from defined groups into a co-design process with new people with similar experiences (Morley et al., 2024). Collaboratively with a sub-group of the Attune Youth Advisory Boards (Batool et al., 2026), we drew upon Normalisation Process Theory (May et al., 2009) and Greenhalgh et al.’s (2004, 2017) guidance on developing and implementing complex interventions to plan co-design workshops (detailed in our protocol, Hugh-Jones et al., 2024). We delivered three workshops, approximately six weeks apart, in each of our three regions, bringing each region’s participants together. Workshops were delivered by three research staff supported by a trauma psychotherapist to support psychological safety. Our approach was informed by our work on relational ethics and trauma-informed practices (Pavarini et al., 2021). Arts-based activities, particularly drawing, collage and drama (role play and improvisation-based on forum theatre, Boal & McBride, 2014), were used to invite playful exploration, to offer emotional distance and to help young people at varying stages of linguistic and cognitive development to contribute on their own terms (Desmond et al., 2015).

The aim and structure of each of the three workshops, and our data collection and approach to analysis, is reported in our protocol paper. In brief, workshops began with a presentation of dominant findings from Attune’s first workstream. The aim of that workstream, which worked with a highly diverse cohort of adolescents (n=74), was to capture their lived experiences of ACEs and what they had felt to be subsequent risk or protective factors influencing their current mental health as adolescents. The methods and findings of that workstream are reported elsewhere (Hugh-Jones et al., 2026). AEBCD workshop one focused on reaching consensus about which aspect(s) of these lived experience findings the co-design participants wanted to address. Outcomes from each region’s workshop were shared in successive workshops in other regions to build collective decision-making, with adolescents’ perspectives and ideas foregrounded. Following consensus, workshops two and three progressed from ideation to production of multiple prototypes, to final realisation of a co-created resource. Implementation and evaluation approaches for the resource were also collaboratively produced.

#### 2.2.3 Outcomes from AEBCD

##### (1) Workshop 1: Consensus aim of the resource

Following the first workshop, participants reached a clear consensus that the primary aim of the co-designed resource should be to strengthen professionals’ capacity to validate young people’s experiences. Invalidation by adults was described by adolescents as pervasive across multiple domains of their lives, with immediate and significant impact on their wellbeing, trust in others and engagement in the setting. Our adolescent co-design participants echoed this. They emphasised that invalidation not only resonated strongly with their lived experiences but also that invalidation underpinned several of other adverse experiences from workstream one. For example, school adversity was often characterised by adults’ dismissal of a young person’s struggle; masking was described as a response to the invalidation of neurodivergent identities; and identity-based abuse was understood as the invalidation of one’s ethnic, gendered, or other important aspects of self. Invalidation was described by adolescents in both Attune workstream one and our co-design study as occurring often multiple times per day in settings and services designed to support young people. They conveyed how invalidation happened in everyday interactions (e.g. in class, in community settings) as well as in high-stakes conversations (e.g. disclosures and asking for support).

Table 2 reports example experiences and categories of invalidation recorded in our workshops, which mirror much of Attune’s workstream one data. Our participants collectively defined invalidation as: *“someone telling you, or otherwise signalling to you, that you are a problem and/or are not important, and that your current experiences and needs are not legitimate or worthy of serious consideration.”*

**Table 2:** Examples of types of invalidation by adults reported by adolescents (n=18) in the AEBCD workshops.

| **Broad categories of invalidation** | **Sub-categories** | **Examples reported in Attune of professionals’ statements or behaviours** |
| --- | --- | --- |
| Dismissing Emotions | Ignoring, minimising or correcting emotions | ‘You’re overreacting’.<br>‘You’re not depressed, you just don’t try’<br>‘There is no need to be anxious’<br>‘You weren’t that close to him, were you?’ (on death of uncle) |
| Rejecting the impact of ACEs | Disbelieving, blaming, comparing, forgetting | ‘It can’t have been that bad. You’re exaggerating’.<br>‘This can’t be an excuse all the time’.<br>‘Other people have it much worse’.<br>‘The bruises don’t look that bad’.<br>Forgetting the adolescents’ past or present adversity (e.g. blaming them for being late despite being a young carer) |
| Ignoring Needs and Perspectives | Ignoring, tokenistic listening, excluding from decision making | Rushing the conversation or talking over them.<br>Not looking at the young person.<br>Changing the subject.<br>Offering quick fixes.<br>Not taking action to help. |
| Misconstruing Behaviour | Pathologising, labelling, focusing on behaviour only | ‘You are being dramatic / attention-seeking’<br>‘You are being thoughtless / selfish / unkind’<br>Responding to sensory overwhelm as ‘difficult’ behaviour<br>Responding to behaviour as ‘bad’ and purposeful<br>(‘Well, you’re not easy are you?’) |
| Invalidating Identity | Discrimination, judging, excluding, ignoring | ‘If you dressed differently, this wouldn’t happen’ (e.g. being bullied)<br>‘You need to change to fit in’ (e.g. mask) |

All co-design adolescents were clear that invalidation by adults constitutes a significant risk to their mental health. Their accounts of this are summarised mechanistically in Table 3. In brief, they explained that invalidation ‘keeps the harm going’ by repeating oppressive and damaging messages and experiences, limiting potential for post-traumatic recovery via relational safety and fostering self-worth.

**Table 3:** AEBCD workshop examples of participant lived experience of the mechanisms by which invalidation by adults harms the mental health of adolescents following ACEs.

| CHILDHOOD |  |  |  | ADOLESCENCE |  |
| --- | --- | --- | --- | --- | --- |
| Dimension of ACE | Harmful message | Adaptive 'behaviour' | How YP try to emerge from the harm | Harmful messages via invalidation by adults / professionals | Continued adaptive 'behaviour' (outcomes) in adolescence |
| Adults using 'power over' them / lack of relational safety | You are not safe with me. I will not protect you. I might harm you. | Hypervigilant to cues of 'power over'. Reticence in relational contact | Reaching out to adults explicitly and implicitly through words and behaviour | You are still not safe. You are not okay. You need to change to be loveable / included / accepted | Anxiety, fear, high arousal / vigilance |
| No support in understanding and legitimising one's own thoughts, feelings and needs | You do not make sense. I know you better than you can know yourself. Your thoughts, feelings and needs are wrong, unimportant, dramatic, attention-seeking, exaggerated, toxic | Minimisation, concealment or abjuration of one's experiences, views, feelings or needs OR intensified to secure a response | Efforts to express their thoughts, feelings and needs. | You still do not make sense (to me or to yourself).<br>I will tell you how you should 'read' yourself.<br>You cannot trust yourself.<br>Your needs, feelings and thoughts are not legitimate here | Low self-worth and shame<br>Confused by, minimising or unable to be informed by one's experiences and needs OR intensifying to secure a response. Possible internalisation and harmful retroflection leading to self-harming behaviours |
| Not being accepted | You are not okay. You need to change to be loveable / included / accepted | People pleasing<br>Denial of self<br>Rebellious position (I will please myself) | Courage to build personal identity | You need to try harder to fit in. You cannot be different | Low self-worth, shame, isolation.<br>Self-denial / lack of self-acceptance |

Importantly, co-design participants emphasised that they recognised professionals’ general intention to be helpful and that their invalidating practices often stemmed from their own stressful and fast-paced working environments, emotional burnout, lack of validation from their employer, colleagues and sometimes adolescents themselves, equating validation with legitimising unacceptable behaviour and / or with an expectations to provide immediate solutions, and not knowing how to be validating.

Given the reported widespread experience of invalidation and its harmful effects, it was chosen by our co-design participants as the problem to tackle via a co-created resource. Participants decided that the opportunity for change lay in first helping professionals to recognise the harms associated with invalidation and to equip them to deliver more validating interactions with adolescents in their practice contexts.

This was the consensus about the purpose of the planned resource.

##### (2) Workshop 2: Priorities for resource structure and content

Workshop two first sought to identify the motivations of professionals for change (i.e. to deliver more validating practices), and what they needed from a resource to help them do this. Reported motivations included: to enjoy their job more by working according to their values for relational care, to get into fewer arguments with young people, to feel able to progress their work with a young person (e.g. social care, employment support), to be less exhausted after a working day and less likely to quit (due to reduced conflict with young people). A further aim of workshop two was to identify priority principles and content of a resource to be created. Pre-defined requirements (i.e. a condition of funding) were that the resource should promote trauma-informed practice, and be low-cost, easy to implement and scalable across diverse public sector settings. Additional requirements specified by our professionals were that the resource should: be different to typical professional training by foregrounding it as youth-led; feel do-able and support quick and practical change; and align with the values of professionals and their organisation/ setting. Co-design adolescents’ additional requirements were that the resource should: include young people as partners in professional training; draw on arts-based methods; and pursue authentic and sustainable change in organisational culture and practice. Adolescents and professionals reached consensus that the resource should also be developmental (e.g., move from awareness of invalidation to supporting change in professionals’ behaviour), use examples of young people’s everyday experiences of invalidation as a catalyst for change and include options for creative approaches to change.

Creative and arts-based approaches were regarded by both adolescent and professional participants as important components of the resource. This was partly influenced by their enjoyment of, and engagement with, the creative methods used during the co-design workshops, such as designing ideal safe spaces and generating improvised characters and stories to explore experiences of (in)validation. Participants felt that creative approaches would appeal to future adolescent users because they are engaging, developmentally flexible, and support agency. In particular, both groups identified forum theatre as a novel and potentially impactful element. Forum theatre is a participatory, drama-based method in which a short scene depicting a problematic or oppressive interaction is co-created, performed, and then replayed. Through embodied perspective-taking and youth-led redirection of the scene, participants can experiment with improved responses (Boal & McBride, 2014).

Participants felt that these pedagogical approaches would help both adolescents and professionals explore difficult experiences and conversations in an emotionally tolerable and potentially empowering way, by creating distance, containment, and opportunities for decentring. Compared with more didactic forms of professional training, they believed creative methods were better able to evoke curiosity, reduce resistance, and foster embodied learning through perspective-taking. For adolescent participants, this was especially important, as they felt meaningful changes in professional practice required practitioners to move beyond intellectual understanding towards an embodied appreciation of the impact of invalidation.

Professional participants endorsed this view and anticipated that experiential learning would be more memorable and relevant to practice than traditional staff development approaches.

Following the delineation of resource principles and priority content, co-design participants ideated, sketched and pitched a provisional resource idea that would satisfy some of the above requirements. This permitted discussion and identification of feasible and engaging components of a potential resource (e.g. easy to start using, not too complex), as well as aspects that may not translate well across diverse settings (e.g. due to time or space constraints or being likely to be seen as too abstract). Between workshops two and three, the research team created five prototype resources based on the ideas produced across the study regions. These were presented for discussion in workshop 3.

##### (3) Finalising the resource content, implementation and evaluation

Workshop three aimed to select and refine core components of the final resource prototype and to identify optimal implementation approaches and priorities for evaluation (data types and data collection methods). Participants opted to merge elements of the prototypes and reached consensus on a three-part resource, described in detail below. In brief, they wanted a resource which enabled the realisation of the prevalence and impact of invalidation in young people’s lives and supported practicing validation, ending with ways to transfer of learning from individuals using the resource into their organisational settings. For implementation, professionals emphasised the need for a strong launch of new resource into their organisations, provision of ongoing support for resource use and sustained senior leadership support. Workshop three also focused on establishing primary and secondary outcomes and suitable evaluation methods. This was extremely complex, as different settings had different capacities for, and interest in, different types of outcome data. For professionals, these included: intention to stay in the job; improved job satisfaction; greater self-efficacy to support adolescents; reduced burnout; and reduced conflict with adolescents. Adolescents’ suggested outcomes included: improved self-assurance; making less risky choices and behaviours; greater self-understanding and trust; improved trust in adults; and hope for the future.

#### 2.2.4 The Final Resource

Via consensus, the final co-designed resource (called Validating Voices) aimed to help public sector professionals and young people (12-24yr), who have lived through ACEs, to work together to embed validation as a helpful experience in settings for young people. Structured as a box of cards, it is a low-cost, low-burden resource that professionals can use flexibly in small groups involving staff and young people in their setting to support the setting to be more validating for young people. Initial cards in the resource present explanations of resource creation, the value of validation for youth mental health, and guidance and options for resource use.

Subsequent cards present ideas for creating psychological safe spaces for young people to engage with the resource with professionals. Following this, the main resource content is structured into three sections, designed to be easy to pick up and use, and which support increasing engagement with the experience of (in)validation and supporting change in professional / organisational practice. The resource is structured to be engaged with sequentially by ‘players’, over whatever time period they choose, but ideally two to three sessions on each section of the resource. It has an accompanying learning log, to be completed briefly and collaboratively by groups at the end of their session. Additional supporting materials, included filmed scenarios and youth co-created serious comics, were made available on the project website (www.attuneproject.com) to support resource use. Table 4 presents the main content sections of the resource with examples.

**Table 4:** Validating Voices key content and examples.

| Section Name | Aim | Materials | How to use | Examples |
| --- | --- | --- | --- | --- |
| 1.REALISE | To help professionals realise the nature, diversity and extent of invalidation experienced by young people (in diverse settings)<br>To give young people a vocabulary and understanding to talk about invalidation<br>To develop professionals' skills for validating practices | • Cards (total 48) colour coded to reflect types of invalidation, spanning invalidation of a young person's: past adversity / trauma, present circumstances; present feelings and needs'; and current identity. Two additional categories are: play / create and reflect.<br>• Cards have a Side A and a Side B. Side A presents a short scenario of an invalidating practice (usually a verbal exchange) between a professional and a young person. *Prompts: 'what do you notice or feel?', 'What might happen in your setting that is like this?', 'Would it help if the adult in the scenario said X rather than Y'?* [a validation skill is suggested] and *'How else could the adult respond in a more helpful way?'*<br>• Side B: a brief trauma history or a current adverse circumstance is revealed about the young person in the scenario. *Prompt: 'does this information make a difference?' and 'what's your take-away learning'.*<br>• Cards for the additional two categories present varied stimuli for discussion, spanning responses to artwork (e.g. depicting safety) to light-hearted prompts (e.g. create the perfect professional; would you rather talk to a meerkat or a gorilla?) | Professionals and young people (6-8 ideally), for approximately 45–60-minute sessions. Can repeat play 2-3 times with the same group, until all cards are explored.<br>Professionals remove any cards in advance that may be unsuitable for the specific players.<br>Players pick any card (could roll a colour coded dice). Players reads Side A, and the group are invited to respond to prompts. Following this, the player is asked to read Side B, and group respond to prompts.<br>Collaboratively note key take-away learning about validating practices for the setting in the learning log. | **Card types: Invalidation of Present Feelings and Needs**<br>**Side A:** Dina is angry. Someone has taken their stuff again. A passing adult sees that they are angry and says "Just calm down. It's no big deal" *What do you notice or feel? Would it help if the adult said "I can see you are really upset. Let's take a minute".* [Validation skill: Noticing]<br>**Side B:** Dina's mum is emotionally abusive. She gets angry when Dina loses stuff and yells at her that she doesn't deserve any new stuff. Dina works hard to take care of her things. She feels nervous at home all the time. And she also feels angry that no-one is noticing. *Does this information make a difference? What's your take-away learning?'*<br>**Card type: Reflect**<br>**Side A:** Here is a picture of a shield created by a young person in Project Attune. *What do you notice? What do you wonder? What would your shield look like?*<br>**Side B:** This shield was created by Sam who has lived through frightening times as a child. Sam feels 'the fear' is always with them. They think that most people are out to get them. They have an invisible shield with them every day. Having a shield can sometimes mean that Sam comes across as weird or unfriendly. *What's your take-away learning?* |
| 2.RE-PLAY | Using curiosity, serious play and embodiment to | • *Set Up: creating safe and validating spaces for RE-PLAY.* Cards guide professionals to practice curiosity, | For use after REALISE cards, if professionals and young people wish to | **Part 1: Improving conversation scripts.**<br>Daniella is in trouble. A teacher thinks she is behind a rumour that has been spreading about |
|  | understand how validating conversations look and feel<br>To motivate and equip professionals to deliver more validating care for young people in their setting | transparency, invitations, playfulness, choice and validation when using the resource<br>• *Part 1: Improving conversation scripts.* A selection of cards showing playscripts of conversations between a young person and a professional. Prompts explore where invalidation happened, its impact and how it could have gone better.<br>• *Part 2: Role-Play.* This has two types of stimuli. First, a series of short (one-minute) filmed exchanges (available on the project website) between an adult and a young person depicting invalidation, and later an improved, more validating exchange. Second, a series of cards inviting players to devise a role-play depicting invalidation | develop more understanding and skills<br>*Part 1:* working in groups of 2-3 for 10 -15 minutes, script writers collaborate to improve the scripted exchange<br>*Part 2:* Film viewers are asked to discuss what they noticed and suggest improvements. They few one option of an improved exchange and discuss. For live role-play, the audience is invited to direct an improved, more validating response from 'the professional'. | another girl. Daniella was not responsible and actually tried to stop it. The teacher addresses the whole class. [script is represented] *Prompts: where are moments of invalidation? Does it make a difference to know that Daniella was abused as a child and that no-one took her seriously? Re-write the scenario to go better for Daniella*<br>**Part 2: Create a role-play**<br>Vishal is a refugee, living in a hostel for three months. They are told to report bad behaviour to an official. Vishal tells an adult official that there is a lot of noise in his room at night that makes him nervous. Sometimes people try his door. The official tells him it is normal for people to have fun at night. Vishal has previously been threatened by older boys and has witnessed someone being stabbed. *Audience prompts: what did they notice? Where did invalidation happen? Ask the actors how it felt? Re-direct for a better outcome* |
| 3.REMEMBER | To capture learning to drive organisational change<br>To use creative, youth-led approaches for organisational change | Draws on learning from REALISE and REMEMBER (learning logs and discussions) to inform creative strategies to support professionals in the setting to delivery more trauma-informed and validating care<br>Cards first establish the importance of translating learning into action which has a high chance of enabling professionals to be more validating.<br>Cards offer ideas for simple, creative youth-led ways to present the learning from the resource to the setting. | A playful, dedicated time and space allocated to being creative about messages, reminders and skills the young people want to encourage in the professionals around them | • Ideas provided: Posters, poems, acronyms, comic strip, a play, create a board game<br>• Big or small messages (e.g. 'Invalidation hurts. Validation helps'; 'Listening to a young person matters to them. I know you are listening to me when you let me speak').<br>• Poster- based on acronym: e.g., WAIT (Wait, Acknowledge, Investigates, Tune in)<br>• Displaying examples of validating statements that professionals could use in the setting |
| LEARNING LOG | To centre the importance of collective learning | Notebook (kept with resource) | Adolescents and professionals together note 2-3 learning points after their use of the resource in that session. Reflect back on this to inform the Remember activities. | • “We learned that not listening properly is the cause of a lot of misunderstanding and upset. We need to think about what listening properly means even when we are rushing”<br>• “Giving a young person choice (e.g. when to meet, their preferred pronoun) helps them to feel recognised” |

The co-designed implementation strategy was (i) a training workshop for staff on resource use, (ii) an opportunity to learn from other settings about how they were using the resource and (iii) support from the research team if needed. However, as per our protocol, we adopted a predominantly ‘let it happen’ approach to learn from organisational agency and creativity in implementation (Duff, 2026). As a minimum, and in line with recommendations for TIA, organisations were encouraged to nominate someone to lead resource implementation in their setting.

### 2.3 Evaluation Stage

#### 2.3.1 Aim

We conducted a preliminary evaluation of the resource aligned with the early stages of resource (intervention) development. We aimed to understand how organisations used the resource, its feasibility, acceptability and in principle efficacy, and how should it be refined for future implementation in the public sector.

#### 2.3.2 Recruitment and Participants

The nine organisations involved in our AEBCD stage were invited to implement the resource in their setting for a minimum of six months. All agreed except for two (the special school and one university student support service) who declined due to low staffing. We extended an invitation to three further settings who had shown interest. These were a coastal community group supporting homeless young people, a coastal town secondary school and a large urban foster care service. These were successfully recruited. Informed consent was obtained by the organisation and any staff member who agreed to be part of the evaluation data collection. One of the original AEBCD settings (a city college) dropped out early in the evaluation stage, citing insufficient capacity to remain involved. A total of nine settings were therefore recruited to the evaluation stage, exceeding our target of six. Table 5 shows the continuation of settings from the co-design to the completion of the evaluation stage.

**Table 5:** The involvement of settings (n=11) across the co-design, implementation and evaluation stages.

| Setting | Co-design Stage | Implementation and Evaluation |  |
| --- | --- | --- | --- |
|  |  | Started | Completed |
| Domestic abuse support organisation | ✓ | ✓ | ✓ |
| LGBTQIA+ community group | ✓ | ✓ | ✓ |
| Care Leaver's Service | ✓ | ✓ | x |
| Group for Underserved Communities 1 | ✓ | ✓ | x |
| Group for Underserved Communities 2 | ✓ | ✓ | x |
| Homelessness Charity | x | ✓ | ✓ |
| University Student Support Service 1 | ✓ | x | x |
| University Student Support Service 1 | ✓ | ✓ | x |
| Secondary school for SEND pupils | ✓ | x | x |
| City College | ✓ | ✓ | x |
| Mainstream secondary school | x | ✓ | ✓ |
| Foster Care Service | x | ✓ | ✓ |

#### 2.3.3. Data collection

Given the lack of suitable UK generated measures suitable for the evaluation of TIA in diverse settings in our study (as discussed in Hugh-Jones et al., 2024), we designed a bespoke three stage evaluation process, informed by participant preferences and priorities in the AEBCD stage. Data collection in this evaluation stage was entirely from professionals who planned to, or did, use the resource in their setting. Stage 1 was a survey administered prior to the resource training session. Ten items asked professionals to self-report their emotional capacity and skills for working in trauma-informed ways with young people and nine items asked respondents to rate their knowledge of ACEs, TIAs and invalidation.

Stage 2 was a 15-item survey completed within two weeks post-training. Items were based on the NPT and relevant domains of Diffusion of Innovation. NPT identifies factors that may promote or inhibit the routine implementation of complex interventions into everyday practice. NPT factors include coherence (of the resource and with the organisation’s aims), cognitive participation (likely commitment and engagement of users), collective action (the impact on routine work, and the work involved) and reflexive monitoring (whether users value the intervention). Included Diffusion of Innovation domains were ‘relative advantage’ and ‘compatibility’ (see Supplementary Materials for all survey items). Stage 3 of the evaluation involved re-administration of the Stage 2 survey, to determine whether NPT factors had influenced implementation success, and semi-structured key informant interviews with any resource users in participating settings. Interviews were conducted by members of the research team and recorded for analysis. They explored organisational decisions for resource use, barriers and solutions to use, observed effects, unanticipated benefits or adverse impact, other lessons learnt and recommendations for improvements, testing and upscaling.

#### 2.3.4 Resource Implementation

A three hour, highly interactive training session was designed by the research team involved in the AEBCD stage (a trauma specialist, an implementation specialist and a participatory arts expert). Training allowed settings to explore and test out each section of the resource and to devise concrete next steps towards resource use within their setting. Participants were invited to join an online workshop at month three to learn how other organisations were using the resource and could contact the research team at any point for support.

#### 2.3.5 Data analysis

Survey responses were summarised descriptively and interviews were analysed using thematic analysis by the lead author.

#### 2.3.5 Findings

##### 2.3.5.1 Quantitative Data

Training was delivered in the nine consented organisations and reached n=49 professionals. Survey 1 was completed by n=26 (53% response rate; Table 7). Given the sectors in which the professionals worked, and the fact that many had been part of the AEBCD stage, it is understandable that they would report a high level of knowledge and capacity to work in trauma-informed ways with an adolescent population with a history of ACEs. A small proportion indicated feeling wary of conversations with young people (item 2), feeling too rushed or stressed to relate well to them (item 3) and feeling uncertain how to respond to them (item 4). Most respondent agreed that validation by professionals may have a significant impact on a young person with ACEs (item 18) although a small number were unsure about what is meant by a validating or invalidating interactions with them (items 13 and 14). Only 4 respondents felt that invalidation would not affect a young person (item 19).

**Table 7:**
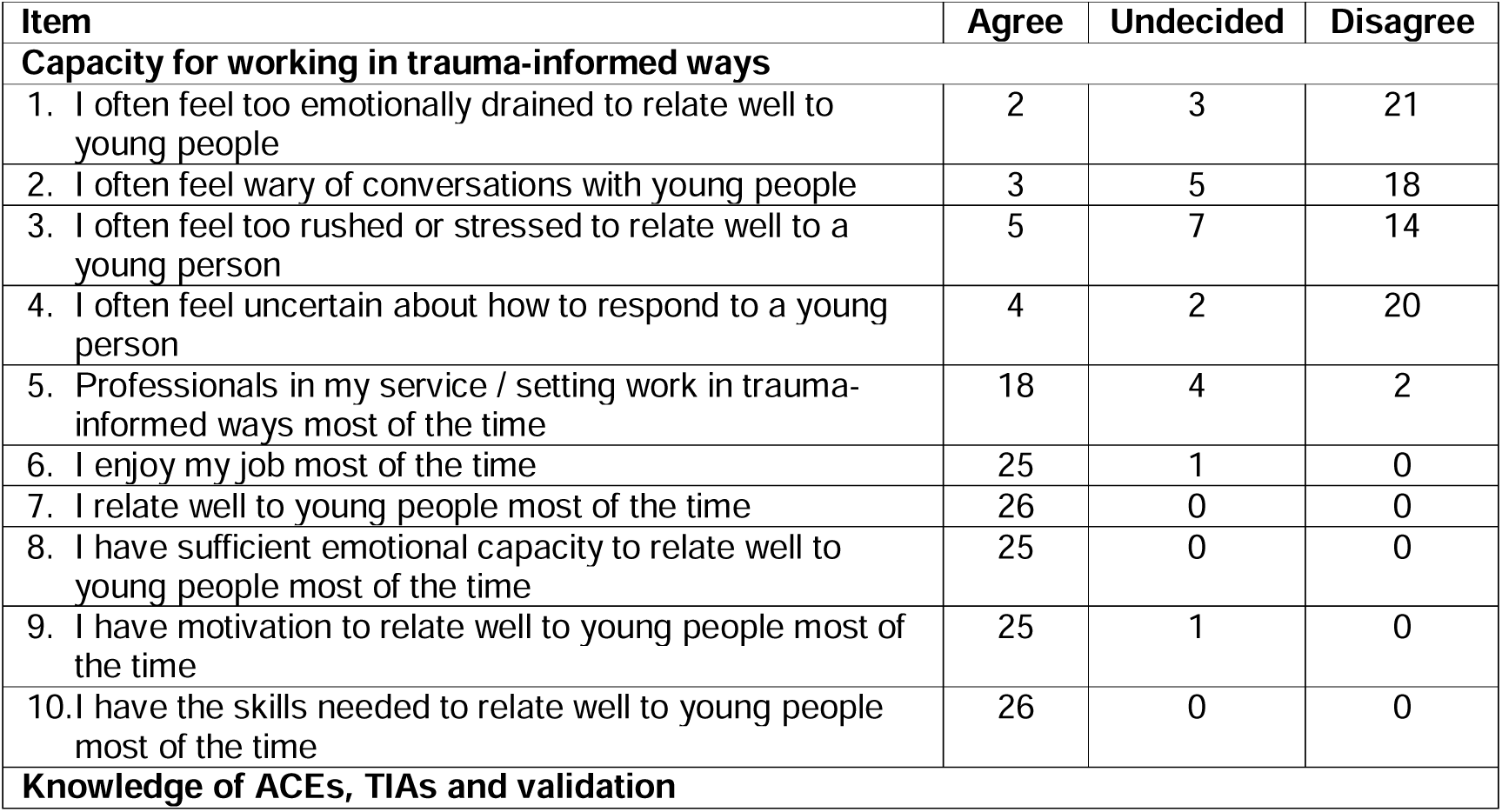
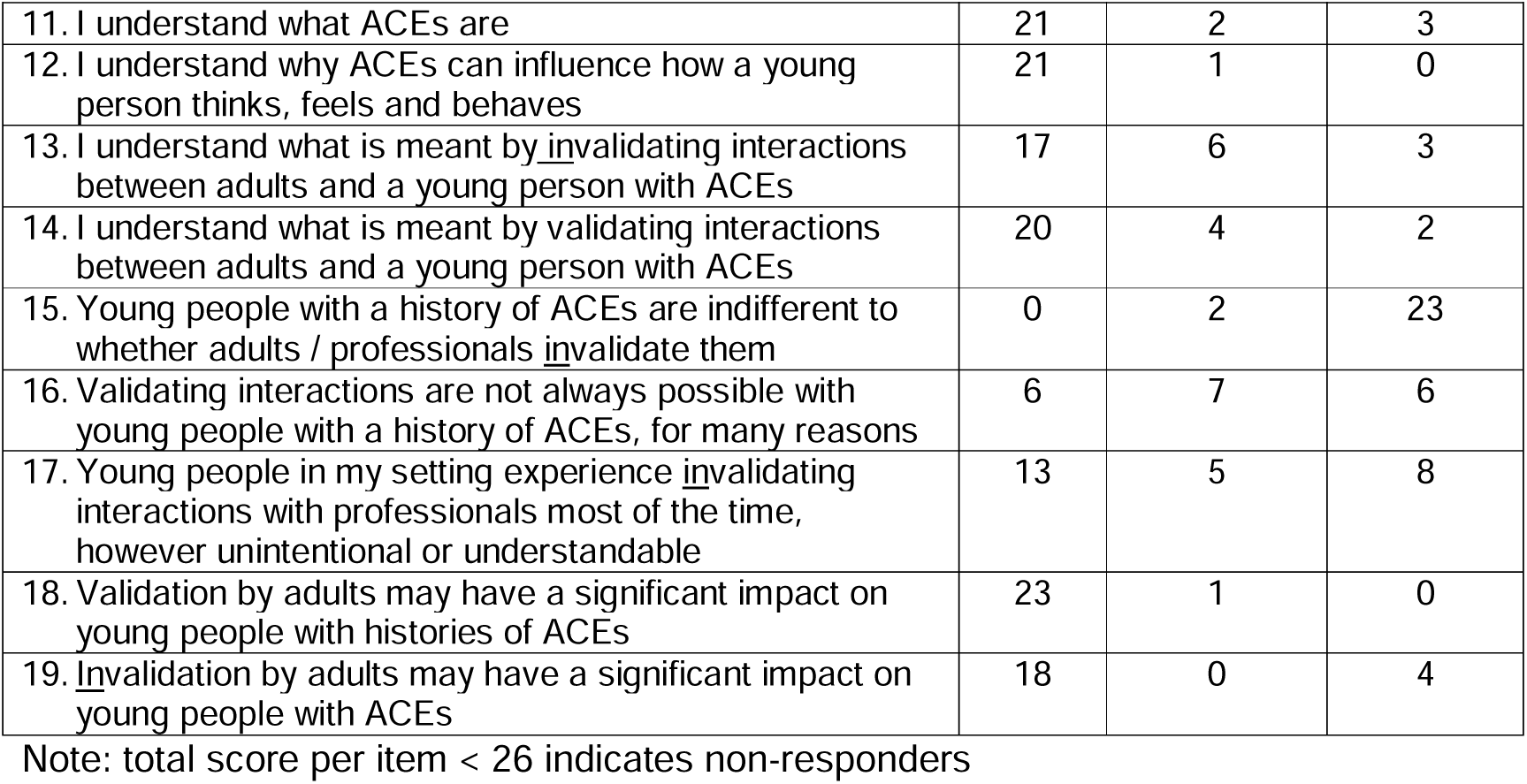
Survey 1 for professionals pre-training (n=26 respondents)

| Item | Agree | Undecided | Disagree |
| --- | --- | --- | --- |
| **Capacity for working in trauma-informed ways** |  |  |  |
| 1. I often feel too emotionally drained to relate well to young people | 2 | 3 | 21 |
| 2. I often feel wary of conversations with young people | 3 | 5 | 18 |
| 3. I often feel too rushed or stressed to relate well to a young person | 5 | 7 | 14 |
| 4. I often feel uncertain about how to respond to a young person | 4 | 2 | 20 |
| 5. Professionals in my service / setting work in trauma-informed ways most of the time | 18 | 4 | 2 |
| 6. I enjoy my job most of the time | 25 | 1 | 0 |
| 7. I relate well to young people most of the time | 26 | 0 | 0 |
| 8. I have sufficient emotional capacity to relate well to young people most of the time | 25 | 0 | 0 |
| 9. I have motivation to relate well to young people most of the time | 25 | 1 | 0 |
| 10. I have the skills needed to relate well to young people most of the time | 26 | 0 | 0 |
| **Knowledge of ACEs, TIAs and validation** |  |  |  |
| 11. I understand what ACEs are | 21 | 2 | 3 |
| 12. I understand why ACEs can influence how a young person thinks, feels and behaves | 21 | 1 | 0 |
| 13. I understand what is meant by <u>invalidating</u> interactions between adults and a young person with ACEs | 17 | 6 | 3 |
| 14. I understand what is meant by validating interactions between adults and a young person with ACEs | 20 | 4 | 2 |
| 15. Young people with a history of ACEs are indifferent to whether adults / professionals <u>invalidate</u> them | 0 | 2 | 23 |
| 16. Validating interactions are not always possible with young people with a history of ACEs, for many reasons | 6 | 7 | 6 |
| 17. Young people in my setting experience <u>invalidating</u> interactions with professionals most of the time, however unintentional or understandable | 13 | 5 | 8 |
| 18. Validation by adults may have a significant impact on young people with histories of ACEs | 23 | 1 | 0 |
| 19. <u>Invalidation</u> by adults may have a significant impact on young people with ACEs | 18 | 0 | 4 |

Note: total score per item < 26 indicates non-responders

Survey 2, administered post-training and before the setting had begun resource implementation, aimed to assess professionals’ initial appraisal of the resource. However, despite multiple reminders, the response rate was very low (n= 5, 10.2% response rate). Of those who did respond, all rated the resource highly and in alignment with their organisational values (see Supplementary Materials for responses). Also despite multiple reminders, Survey 3 attracted only one respondent and therefore no data is reported here.

##### 2.3.5.2 Qualitative Data

Seven key informant interviews were conducted across the five organisations that had trialled the resource and were available for interview. Three of the recruited nine settings reported being unable to trial the resource; for two community organisations this was because of staff unavailability to lead this, and in the care-leaver service this was due to staff changes and their preference to wait until they had introduced a coherent suite of professional development activities. Across the five settings who reported on resource use, implementation varied considerably, and from its anticipated use. As summarised in Table 8, the resource was implemented with staff and young people, and in groups / classes, one-to-one conversations, in staff reflective practice, in carer discussions, through creative activities, through peer facilitation, and as part of broader participation and relationship-building work.

**Table 8:**
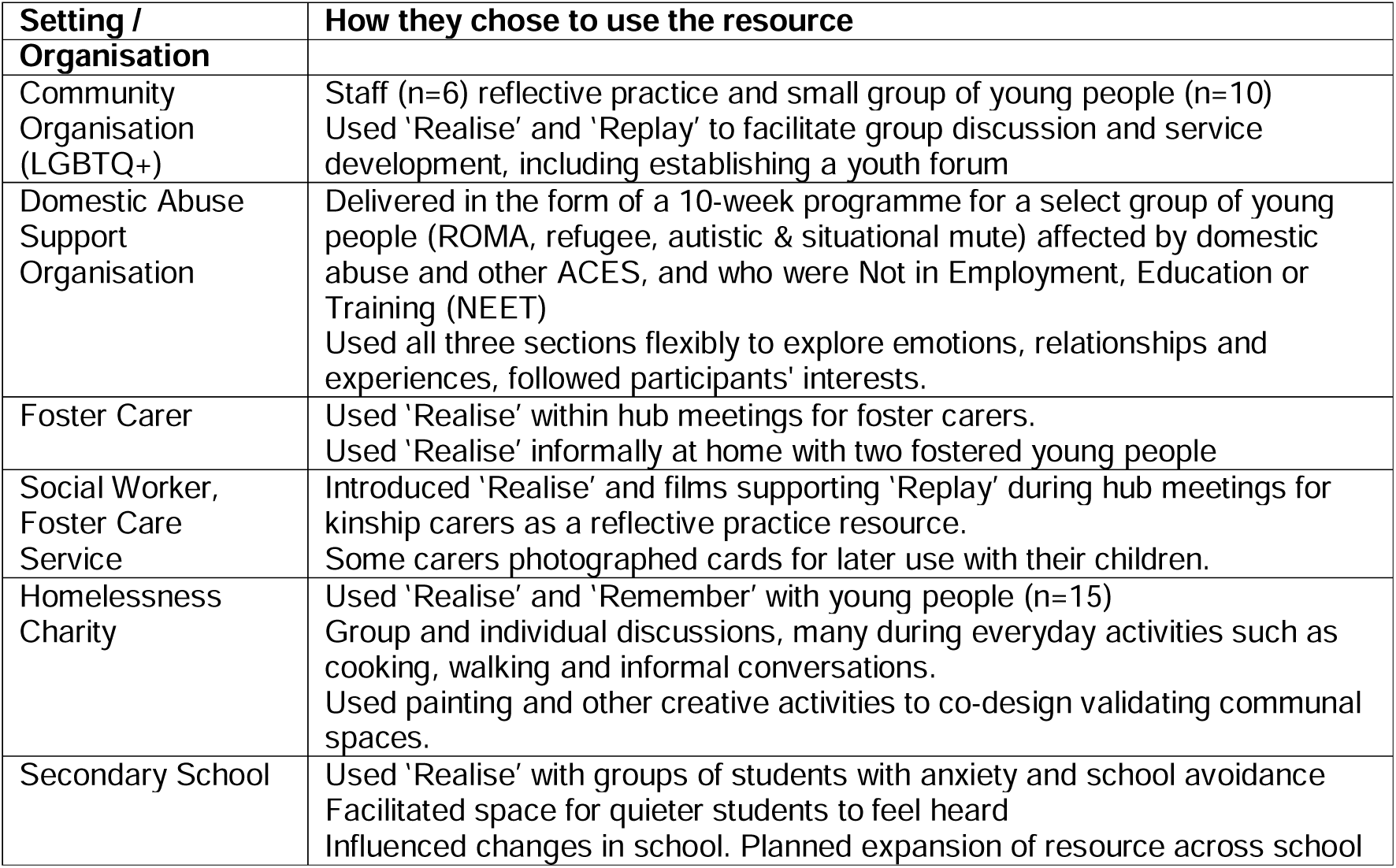
Summary of implementation choices by participating organisations.

Thematic analysis identified five themes relating to the feasibility (Theme 1), perceived effects and mechanisms of action (Themes 2-5) and future development of the resource (Theme 5).

Theme 1 Flexibility, creativity and youth-led design supported resource feasibility

Across settings, participants mostly described the resource as acceptable and feasible to implement. Acceptability was supported as the resource aligned with organisational values, particularly commitments to relational practice, amplifying young people’s voices in service development and promoting trauma-informed approaches: “We *want to make sure that adults and professionals and people who are providing the services know what it is that they [young people] want…the resources was very much aligned with our approach”* (Domestic Abuse Support Organisation). Similarly, the foster care service viewed the resource as fitting naturally within existing reflective practices involving social workers and carers, *“so straight away you could see it being a really good learning tool.”*

A notable finding was the extent to which organisations adapted the resource for local use. Although originally designed as a structured process through which young people and professionals could collaboratively develop validating practices within that setting, no setting implemented it exactly as envisaged. Instead, organisations incorporated resource components into existing activities and relationships (Table 8) and participants reported this adaptability as a strength and reason for feasibility.

Participants also attributed acceptability to the fact that the resource content (i.e. scenarios on cards and films) were highly relevance to the lives of young people, and as sufficiently open to accommodate diverse experiences and developmental stages: *“they can be tailored to different backgrounds and where people have come from or what stage of their journey they’re at with us” (Homelessness Charity).* For some settings, implementation moved beyond formal sessions and became embedded within everyday practice. The homelessness charity described spillover effects into everyday conversations to the point “*where we use it as part of normality - they’re [young people] like, oh, are we getting that box out today? And it’s not all been let’s sit down and talk about this. It’s…have a conversation walking or driving with a client.”* Adaptations also included using the Realise cards for targeted support to young people who were “*drowning*” and at risk of exclusion (Secondary School) or who were *“about to fall off that cliff edge of support”* (Domestic Abuse Support Organisation), as well as for staff-only reflective practice (described below).

Participants also attributed acceptability to resource credibility based in its co-production with young people. While this was noted more often in training sessions than in the interviews, several settings stated this enabled resource engagement to be framed not as ‘just’ another professional initiative, but as an opportunity to learn directly from young people with lived experience: “*it’s thanks to you for building it with young people and listening and having, you know, their perspective. I think it does make a difference. That is a major selling point to our young people.”* (Homelessness Charity)

Theme 2 *The resource helped make hidden experiences visible*

In terms of observed effects, rather than functioning primarily as an educational resource, participants consistently described the resource as helping young people communicate experiences, feelings and needs that were often difficult to articulate.

In so doing, it appeared to create opportunities for experiences of adversity, silencing, invalidation and emotional distress to become visible: *“she’s never really had much practice at saying her feelings and having a language for that”* (Foster Carer). For the secondary school, the resource provided a means of reaching students whose experiences were often overlooked, namely those with emotionally based school avoidance: *“One of the gaps we identified was that we do not hear the voice of our anxious students, of which there are a significant number. This is for several reasons; they are too anxious to speak to adults, they don’t like to speak out loud in front of others, their attendance is often poor…using the resource with these groups gave an opportunity for something different to happen.*” (Teacher 1).

For most settings, participants suggested that these effects were less attributable to the resource materials themselves than to the conditions they helped create.

Creativity appeared particularly important. Through role-play, artwork, storytelling and imaginative activities, young people were able to explore experiences indirectly rather than through direct disclosure. Professionals described these approaches as youth-friendly and trauma-informed, enabling experiences that had previously remained hidden, misunderstood or difficult to express to become visible. In two settings, young people were described as *"really into it"* (Domestic Abuse Organisation) and *"so passionate about it"* (LGBTQ+ Community Organisation) when engaging with creative elements of the resource. The homelessness charity similarly reported that creative activities supported self-expression, validation and belonging, noting that displaying artwork produced through the Remember activities had *"changed the whole dynamic of the hostel",* with young people taking pride in showing visitors work that reflected their experiences.

Across settings, resource use was described by participants as supporting a recurring process whereby young people recognised their own experiences in the stories and scenarios presented within the resource. This recognition often prompted disclosure, discussion and shared understanding in the small groups using the resource: *"Immediately everybody in the group, ’oh yeah, that’s happened to me*.

*That’s happened to me. That’s happened to me"* (Domestic Abuse Organisation). Participants described these moments of shared experience as particularly important because they recognised rather than invalidated young people’s responses to adversity and reduced their feelings of shame and isolation. The secondary school similarly reported that trust and connection through hearing each other’s experiences: *"Trust was built, not just between us and them, but also with each other as peers."* Staff observed that this process helped students realise *"they weren’t on their own and the sense of unity and trust within the group was really very special."* (Teacher 2)

Several participants suggested that psychological safety was central to the emergence of these processes, facilitating conversations that might otherwise not have occurred. Establishing and maintaining psychological safety was a focus of the resource, but professionals implementing this resource were also often trusted adults for the young people. Nonetheless, after resource use, one organisation described young people beginning to share experiences with one another that had remained previously undisclosed: *"Clients that we have struggled with for a long time now want to get involved and then they’re opening up to each other. It opened doors to conversations that haven’t happened, but that we didn’t realise weren’t happening."* (Homelessness charity). For that setting, resource use was also credited with enabling a new safeguarding disclosure by a young person*“that social workers haven’t been aware of that hasn’t been disclosed previously, and I do think it’s [resource use] really made a difference.”*

Theme 3 *The resource supported processing of trauma and invalidation*

Perhaps the most unexpected finding concerned the Replay component. While the activity was originally intended to support professionals’ learning about validation, participants in two settings described young people appropriating the activity for their own purposes. Rather than teaching adults, young people used role-play to revisit, make sense of and rework experiences of invalidation. In these settings, Replay appeared to operate less as a training exercise and more as a vehicle for therapeutic reflection and processing. Two settings in particular observed this. Both described how the invitation to role-play was possible as their groups of young people knew each other and had a level of relational safety, and because they had a member of staff who was a trusted and skilled facilitator. Rather than using Replay as intended (with professionals learning from young people about how to be more validating), young people in these settings used role-play to re-enact and process their experiences of invalidation for therapeutic benefit. The domestic abuse organisation reported that *“by doing that [role-play], that processed something”.* Professionals observed that this process helped challenge adolescents’ self-blame and the belief that their experiences of invalidation were somehow deserved. Reflecting on young people’s responses, one professional commented: “*to know that you’re not alone, I’m not just this person who has all these horrible things happen to them. It’s empowering for them because they are just sitting with this thing that happened to them. It is a transformative tool, isn’t it? It means that they are doing something positive with what has happened to them.”*

Similarly, the LGBTQ+ community organisation described how role-play enabled young people to explore *“moments of kind of micro invalidation*” based on their identity that they experience *“almost constantly.”*. Settings observed that use of Replay wa*s “really quite profound - almost like act it out and process it a little bit…especially those encounters in which there had been a profound sense of invalidation."* Having witnesses to their invalidation (the Replay peer ‘audience’), and within a relationally safe group, was felt to be pivotal to the catalysing of this therapeutic effect as *“the group itself feels quite containing – a type of co-regulation”.* More as intended, the use of Replay in that setting then progressed to what the service, or other professionals in other settings, could do differently to deliver more trauma-informed and validating interactions.

Theme 4 *The resource supported professionals to listen differently*

Across settings, participants reported changes in how professionals interpreted and responded to young people’s behaviour. Rather than focusing primarily on behaviour management or practical problem-solving, professionals described becoming increasingly curious about the experiences, emotions and histories underpinning young people’s actions: *“Staff are listening in a different way than ordinarily or previously. A support worker in our team, she’ll openly say that to me that she is having a different conversation with young people now”* (Homelessness charity). The shift was often described as moving beyond what young people were saying or doing towards exploring the reasons behind those behaviours. Participants repeatedly referred to "reading between the lines", asking different questions and seeking to understand the impact of adversity on young people’s experiences. The homelessness charity provided one of the clearest examples of this shift. Following changes in listening practices, staff reported increased engagement from young people and the disclosure of previously unknown traumatic experiences. One young person reportedly explained why they had disclosed information to a support worker but not their social worker: *"Because they don’t listen."*

The LGBTQ+ organisation described a similar process among staff using the Realise cards during reflective practice sessions. The Side B information, revealing the adversities experienced by the fictional young person, prompted staff to reflect on how their interpretations changed when additional context was provided: “it *was such an interesting way to really dig into kind of how we encounter young people, like within the context of kind of therapeutic support work we offer*”. Participants described these discussions as deepening understanding of validation and helping staff consider how services might respond differently to young people’s distress: *“it was just so helpful to have these different ways of thinking about validation and what adults and professionals and practitioners can do differently and what that might look like … often I think services will kind of do what they need to do to make the service feel good about itself, but not necessarily what the young people need.”*

Perhaps the clearest example of practice change emerged within the secondary school. Listening to students experiencing anxiety and school avoidance led staff to implement a series of practical changes, including more supportive responses to lateness, advance warning of classroom changes and greater attention to the impact of raised voices. Staff reported that visible action in response to students’ concerns led young people to feel they were taken seriously: *“visible changes happening, means that students feel heard and empowered, where they’ve often felt invisible and marginalised due to their anxiety and this has been very validating for them”* (Teacher 1). Teachers reported improvements in attendance, confidence and belonging among participating students, and planned to extend implementation across Key Stage 3 for 900 students through the Personal, Social, Health and Economic program.

Theme 5 Future development and implementation support

Participants identified two main areas for future development. First, several settings suggested that the resource would benefit from additional guidance on preparing young people for participation and establishing the relational foundations necessary for meaningful engagement. Participants emphasised that young people’s readiness to discuss adversity varied significantly, and that trust and psychological safety often required active development before deeper activities could occur. Whilst the resource does emphasise the importance of these, specific strategies for operationalising them were needed: “*maybe kind of work on doing some relationship building first over a couple of sessions or kind of mind map some ideas around the different concepts we might encounter, like what does it mean to have an adult that’s attuned?” What does it mean to experience invalidation”* (LGBTQ+ community organisation).

Second, participants highlighted the need for additional support in implementing Replay. While highly valued by the two settings that used it, other professionals reported lacking confidence in facilitating role-play and were uncertain about how to engage more hesitant young people. The resource recognises the challenges of role-play and included alternative and lower-stakes versions, including re-writing scripts and re-directing the film clips. Some settings had forgotten or overlooked these resources. Nonetheless, the importance of a skilled member of staff was underlined*: “You need somebody there to provide that safety and containment."* (LGBTQ+ community organisation).

Finally, participants highlighted wider organisational barriers to implementation, including staffing changes and competing priorities. While enthusiasm for the resource was generally high, participants recognised that broader implementation in the public sector settings that are busy, with fluidity in people in those settings. For example, the foster care service social worker valued the resource, but saw challenges in implementing it in a structured way, due to varied staffing and carer attendance at hub meetings. Wider implementation across services was felt by the homelessness charity to require word-of-mouth, as they felt personal recommendations of good resources reflected the reality of wider use. The domestic abuse support setting felt that, whilst they valued and used the resource, its wider implementation in schools, for example, would face significant challenges: *“it’s very, very difficult because of the attitude, the attitude barrier to breakthrough with schools… Again, this this culture of invalidation. But there’s a real resistance to changing that.”*

## 3.0 Discussion

Resources, informed by the experiences and needs of young people, are needed to support settings and services working with young people to operationalise TI principles. This study was informed by a large lived experience dataset from Project Attune, which involved very diverse adolescents with ACEs (Hugh-Jones et al., 2026). Working with professionals from public sector settings, and some of their young people, we responded to lived experience data and co-designed a new resource (Validating Voices). Redressing invalidation was the focus of the resource chosen by participants, and the resource aim was to bring public sector professionals and young people into collaboration to help their setting deliver more validating conversations and practices to young people. The resource was implemented in nine diverse public sector settings in England. We discuss key findings from the co-design and preliminary implementation evaluation stage, before considering implications for public sector TIA more widely.

### 3.1 AEBCD: what participants wanted

ACE frameworks largely focus on the correlates of exposure to adversity (Jia & Lubetkin, 2024), but the young people in our study emphasised that what happens after ACEs, particularly their current experiences, also matters. Echoing and extending findings from the early Attune lived experience workstream (Hugh-Jones et al., 2026), they highlighted the harms to mental health and wellbeing caused by professional invalidation. Elsewhere, we offer a mechanistic account of how invalidation affects mental health in ACE-exposed adolescent populations (Hugh-Jones et al., 2026). While invalidation can be harmful for anyone, the power imbalance between professionals and young people, combined with the violation of expected care, appears to make it especially disempowering, confusing, and damaging for adolescents, potentially further heightened for those who have previously known abuses of power in ACEs. Our findings align with evidence from multiple contexts reporting youth experiences of invalidation and how these can lead to avoidance and distrust, increased masking or closedness, and social disconnection (Bansema et al., 2026; Cunningham et al., 2024; Wasson-Simpson et al., 2022).

A striking finding from our co-design stage was that both young people and professionals favoured a relational approach to addressing invalidation. More conventional professional development resources, such as training modules or organisational tools for trauma-informed practice, generated little interest. This preference suggested that invalidation should be addressed at its source by improving relationship quality, respect and empathy and in ways that are attentive to subtle if not overt misuses of power.

Co-designers wanted to foreground the therapeutic value of positive professional encounters, recognising the strengths of professionals to do this, and in which adolescents feel heard, understood, and taken seriously. Resource content therefore focused on developing skills for validation that reflected key therapeutic relational qualities, including openness, listening, curiosity, empathy, understanding, and helpful action (Reis, 2017). Importantly, professionals shared this aspiration for the impact of the resource, expressing their desire to provide these kinds of validating interactions to benefit young people and for their own professional satisfaction and wellbeing.

Co-design participants made some highly innovative choices in resource design. They prioritised a playful, creative approach in resource content. They anticipated this would enhance adolescent engagement and agency, provide an accessible route into discussing invalidation, and create a distinctive, embodied, and memorable learning experience capable of influencing professional practice. They also designed the resource to include art-based methods to support youth voice and creativity in diffusing the learning about how to deliver more validating practices throughout the setting or organisation. Adolescents viewed the resource section of Replay (drawing on role-play) as essential for helping professionals understand the experience and impact of invalidation, and to provide adolescents with power to teach (re-direct) professionals towards more helpful responses in the scenarios.

These design choices were highly novel, as most trauma-informed approaches rely primarily on experts teaching staff via standard pedagogies; whilst these improve staff knowledge, attitudes, and behaviours, evidence of sustained impact remains limited in some contexts, and they often exclude service users as experts (Purtle, 2020).

Whether role-play should be included was discussed extensively during co-design. Participants recognised that the term can be off-putting, that many people would be reluctant to participate, but that others would highly value this form of learning. To address this, the resource adopted a stepped approach, including scripts and filmed scenarios, alongside multiple modes of engagement, such as observer, note-taker, director, or actor. Their choices are in line with evidence which shows that drama-based learning can be impactful, memorable, and effective in fostering empathy and perspective-taking (Wasylko & Stickley, 2023). Studies also show that the benefits of role-play extend beyond active participation, with observers also reporting gains in understanding and professional learning (Rønning & Bjørkly, 2019).

Finally, participants asked for a ‘help it happen’ approach to implementation (Duff, 2026). Professionals strongly advocated for a dedicated launch workshop delivered by the research team to support organisational implementation. They argued that engagement from external experts would confer legitimacy and credibility on the resource, signalling both its professional value and the research team’s commitment to its adoption. Such involvement was also seen as important for generating momentum, establishing shared implementation goals and processes, and fostering accountability for sustained use within the organisation.

### 3.2 Evaluation Stage

The evaluation sought to understand the acceptability of the resource and how diverse settings chose to implement *Validating Voices* during the six-month study period. Given the early stage of resource development, the focus was on implementation processes rather than later-stage outcomes such as adoption, fidelity, or sustainability (Proctor et al., 2011). Five of the nine recruited settings were retained and implemented the resource in some form. These settings spanned social care, education, and the third sector. Qualitative process evaluations are recommended for understanding intervention implementation in complex real-world settings such as these (McGill et al., 2020). Accordingly, our evaluation drew primarily on in-depth interviews with key informants from participating organisations. Despite repeated efforts, we were unable to obtain meaningful quantitative evaluation data through pre-, post-, and follow-up staff surveys. Although the reasons for non-response are unclear, they likely reflect competing professional demands and the perception of surveys as low-priority activities.

### Learning about implementation

Among those who completed the baseline survey, self-reported knowledge of ACEs, trauma-informed practice, and, in most cases, the negative impacts of invalidation on young people were high. As such, findings regarding implementation should be interpreted as emerging from a workforce already oriented towards youth adversity and TIAs.

Local adaptation was central to implementation success. No setting implemented the resource exactly as envisaged, even by those involved in its development. Rather than convening groups of young people and professionals to work systematically through the resource over time, settings adapted implementation to fit their needs, time availability and young people. Such adaptations to interventions are common in practice and can identify creative and unexpected uses and solutions in the field, which can be translated into implementation guidance in future iterations (Kirk et al., 2021; Moore et al., 2013).

Adaptations by our participants were entirely related to resource delivery rather than content and were typically driven by one or two committed staff members. Such commitment was also evident in the three settings that had not participated in co-design, suggesting that prior investment in resource production was not the sole factor underscoring their enthusiasm. Staff members reported drawing on their knowledge of their organisational culture and existing relationships with colleagues and young people to identify feasible ways to use the resource. The importance of such champions for implementation and local adaptation is well established (Miech et al., 2018; Proctor et al., 2023). Champions tend to have particular attributes, capacity and organisational positions to enable implementation (Bonawitz et al., 2020; Santos et al., 2022). However, reliance on champions can concentrate responsibility within individuals rather than embedding new practices within organisational systems more broadly (Stark & Page, 2026). Consistent with this, the three sites that did not attempt to implement the resource reported either insufficient staff capacity to drive implementation (two community organisations serving underserved populations) or difficulties identifying viable implementation pathways within complex organisational structures (a university student support services). This is not an uncommon finding in intervention studies; many settings report failing to implement interventions not because of intervention need or acceptability, but because they lack the resources or expertise to deliver an intervention as intended (Bell et al., 2010).

Importantly, local adaptations by our participants appeared to retain some fidelity of function, if not fidelity of form, i.e. the anticipated active ingredients and mechanisms of change (Hawe et al., 2008; Jolles et al. 2019). Although these were only lightly provisionally theorised in AEBCD, realising the impact of invalidation appeared retained as an active ingredient in all local adaptations. However, less retained was the fidelity of function anticipated via the role-play, where young people would have ‘power’ to inform professional practice and behaviour change.

### Learning about resource use

The aspirations made explicit in the co-design stage for the processes and impact of the resource were partly observed in the evaluation. Settings reported that the creative approaches in Validating Voices supported engagement by some of their young people. Many also noted that young people seemed to respond well to the indirect and symbolic ways of communicating experiences of invalidation, sometimes talking more than in routine conversations with professionals. Shared experiences of adversity and invalidation appeared to legitimise their distress responses, strengthen peer solidarity and support further relational safety in the group.

However, the aspirations of the co-design team regarding the Replay element of the resource were not met in practice. Although some settings reported watching some of short film scenarios within this section, no setting reported professional’s engaging in live role-play in order to ‘experience’ invalidation and be guided by young people on how to be more validating. In three settings (school, homelessness charity and foster care service) the Replay component was felt to be unattractive to users, or as requiring skilled facilitators. Many settings did not have a youth panel via which they could easily cohere groups. More positively, in two settings, young people appropriated the role-play for personal ‘processing’ of invalidation. However, such benefits were only reported in what can be described as psychologically safe groups, with trusted facilitators known to the young people. Thus, whilst the role-play element appeared extremely important to the co-design participants, it may be hard to implement at scale.

### Impact on professional practice

Across settings, respondents reported that the resource supported the visibility of young people’s experiences, fostered relational safety and reflection, encouraged more curious and validating responses from professionals, and created opportunities for meaningful dialogue and disclosure about adversity and its impacts. A consistently reported change was that professionals felt they were listening differently, described as moving away from adult-centric perspectives and solutions to curiosity (i.e. wanting to know more about, and take seriously, the young person’s experience and needs). Side B of the Realise cards, which introduced an ACE back story to the young person in the Side A scenario, was helpful in some settings. For example, foster carers and social workers appreciated being reminded of that every young person has a Side B despite working with young people with ACEs all of the time. Nonetheless, this feature was not commented on as much as expected in the interviews, although they were highly valued in the training sessions. Reasons for this are unclear. This was a distinct attempt to help professionals bring a young person’s past or current ACEs into their understanding of the young person’s present engagement with them.

### 3.2 Strengths and Limitations

Study strengths include building on a large data set of diverse youth experiences on ACEs, trauma and mental health in AEBCD (Hugh-Jones et al., 2026). Our focus on creating tangible resources for the public sector meets a real-world need. Co-design with young people, and centering their experiences and needs, reflects efforts to redress the omission of youth voice in the development of trauma-informed understanding, practices and resources. Diverse young people and setting were involved in both study stages, including those often hard to reach (e.g. homelessness and foster care sectors). Following our protocol (Hugh-Jones et al., 2024), we used key frameworks and process evaluations to guide resource development, implementation and evaluation in real-world, complex settings (May, 2009). Findings contribute to the call for evidence about how trauma-informed practice can work ‘in the field’ (Goldstein et al., 2024).

Findings should be considered in the context of several limitations. Although our AEBCD participants were diverse, sample sizes in line with similar studies (Bielinska et al., 2022), and the resource landed well with diverse young people who recognise their experiences within the resource, the complexity of public sector settings will require engagement with a much higher number of settings to refine our resource.

Survey response rates were very poor and our evaluation rests on the experiences of professionals likely to be positively biased given either investment in resource co-design, or by opting in to trialling the resource.

### 3.3 Implications and future directions

Giving young people space to design resources for professionals is rare in trauma-informed work. Our study showed that young people hope that trauma-informed practice will include attention to the importance of validation in everyday encounters with them. Validation skills may therefore offer a concrete focus for workforce development and a means of operationalising core TIAs. Across settings, resource use supported safety, trust, collaboration and choice. Professionals frequently described "listening differently", characterised by greater curiosity, fewer assumptions, and reduced reliance on adult-centred interpretations or solutions.

Viewed in this way, validation does not need to be considered a specialised therapeutic technique but a set of relational practices that anyone can use.

Whilst our AEBCD stage aligned with early intervention development processes (Wight et al., 2016), future refinement of the resource could draw more explicitly on behaviour change theory and behaviour change techniques to strengthen its impact on professional practice (Michie et al., 2011; 2013). The role-play component, in particular, warrants further development. More broadly, although individual practice change is important, trauma-informed approaches ultimately seek organisation-wide transformation (Rich et al., 2025). Early testing of Validating Voices suggests potential to support youth contributions to organisational trauma-informed development, but this remains provisional and requires evaluation through resource development and piloting. Future iterations could include more explicit support for organisational change and youth-professional collaboration. Notably, our findings suggest that professionals may be more comfortable learning about young people’s experiences than creating structures through which young people can directly influence professional behaviour and organisational practice. Future research should therefore examine how meaningful youth participation can be sustained within trauma-informed systems, using more robust evaluation methods, including realist approaches, to understand what works, for whom, and under what circumstances (d’Souza et al., 2021).

### 3.4 Conclusions

Although preliminary and based on a small-scale implementation evaluation, this work provides an empirically grounded and youth-informed contribution to the trauma-informed field. Our study demonstrates the value of involving young people directly in defining what trauma-informed practice should look like in everyday public-sector settings. Findings suggest that validation may represent a tangible mechanism through which trauma-informed principles such as safety, trust, collaboration and empowerment can be operationalised in routine practice. The findings also demonstrate the potential value of youth-led and creative approaches to implementation, while highlighting the challenges of embedding more ambitious forms of youth influence and power-sharing within organisational systems. Future research should focus on refining the intervention, testing implementation strategies that do not rely heavily on local champions, and examining whether improvements in validating professional practices lead to measurable benefits for young people’s mental health, engagement and longer-term recovery following adversity.

## Data Availability

All data produced in the present study are available upon reasonable request to the authors

## Acknowledgements

We thank the young people and professionals who participated in this research. We acknowledge the UKRI funding (MR/W002183/1) that supported this work through the MRC/AHRC/ESRC Adolescence, Mental Health and the Developing Mind Programme. We thank the numerous community organisations that facilitated recruitment and data collection, and the ATTUNE Young People’s Advisory Board for their guidance throughout the project. We acknowledge all ATTUNE team members who contributed to data collection and analysis across multiple sites and disciplines.

